# Neonatal Hypothermia at and after Admission: Burden and Associations with Outside Air Temperature and Neonatal Ward Temperature in Four Sub-Saharan African Countries Implementing with the NEST360 Alliance

**DOI:** 10.64898/2026.07.04.26357151

**Authors:** Melissa Mar, Hannah Mwaniki, Edith Gicheha, Lucas Malla, Vincent Otieno Ochieng, George Okello, Samuel Ngwala, Opeyemi Odedere, Mariam Thabit Johari, Catherine Paul, Kristina Shemwell, Joy E Lawn, Nahya Salim, Msandeni Chiume, Veronica Chinyere Ezeaka, William M. Macharia, Rebecca Richards-Kortum, Maria Oden, Lisa R Hirschhorn, Natasha R Rhoda, Elizabeth M Molyneux, John Wainaina, Christine A. Bohne

**Affiliations:** Rice360 Institute for Global Health Technologies, Rice University, Texas, USA; Maternal, Adolescent, Reproductive, & Child Health (MARCH) Centre, London School of Hygiene and Tropical Medicine, London, UK; Department of Paediatrics, Aga Khan University, Nairobi, Kenya; Department of Paediatrics and Child Health, Muhimbili University of Health and Allied Sciences, Dar Es Salaam, Tanzania; School of Global and Public Health, Kamuzu University of Health Sciences, Blantyre, Malawi; Ifakara Health Institute, Dar Es Salaam, Tanzania; Ministry of Health, Malawi – Reproductive Health Department; Department of Paediatrics, College of Medicine, University of Lagos, Lagos, Nigeria; Department of Paediatrics, Kamuzu University of Health Sciences, Blantyre, Malawi; Children’s Institute, Department of Paediatrics and Child Health, Faculty of Health Sciences, University of Cape Town, South Africa; Northwestern University Feinberg School of Medicine, Illinois, USA

**Author notes:** Corresponding author: Melissa Mar. With the NEST360 Neonatal Inpatient Dataset and Data Systems Collaborative Group and Context Tracker. Joint first (Melissa Mar, Hannah Mwaniki). joint senior (John Wainaina, Christine A. Bohne).

**Keywords:** neonatal, newborn, hypothermia, thermal care, warm chain, low- and middle-income countries

## Abstract

**Background:** Annually, 2.3 million newborns die, largely from preventable causes. Neonatal hypothermia is an important contributor to morbidity and mortality, particularly in low-resource settings. This study quantified the burden of hypothermia at and after admission in four NEST360-supported countries and examined associations between outside air temperature, ward temperature, and neonatal hypothermia.

**Methods:** We conducted a retrospective analysis of newborn admissions (January 2021 to June 2025) across 66 neonatal units in Kenya, Malawi, Nigeria, and Tanzania. Hypothermia was defined using WHO thresholds (mild: 36.0–36.4°C; moderate: 32.0–35.9°C; severe: <32.0°C). Newborn admission and lowest after admission body temperatures were extracted from routine clinical records. Ward temperatures were captured using the Hadli™ Monitoring System, and environmental temperatures were obtained from Open-Meteo. Multivariate ordinal logistic regression assessed associations between air temperature, ward temperature, and hypothermia at admission and during admission.

**Results:** Among 418,458 newborn admissions with recorded admission temperatures, 47.3% (n=220,684) were hypothermic at admission (country range: 22.8%–61.9%), while 63.5% (n=48,746) experienced hypothermia during hospital stay (country range: 18.5%–74.4%), based on 76,855 admissions (July 2024–June 2025) with temperature data. Based on admission and subsequent temperature, 28.5% had no documented hypothermia, 8.6% improved to non-hypothermic status, 29.4% developed hypothermia after admission, and 33.5% experienced hypothermia at admission and during hospital stay. Across 59 neonatal units, minimum ward temperatures >26°C were maintained on 92.6% of 365 days. At admission, ward temperatures of 30–33°C were associated with 9% lower odds of a lower thermal category versus 26–28°C (p<0.01). After admission, ward temperatures of 28–30°C reduced odds by 18% (p<0.05). Warmer outside temperatures (>24°C day, >21°C night) were protective, corresponding to 19% and 68% lower odds of a lower thermal category after admission, respectively, compared with 19–24°C and 15–21°C reference groups. Newborns had 3.6-fold higher odds of hypothermia at night than during the day. Each 1°C increase in post-admission temperature reduced odds of death by 6%.

**Conclusion:** Neonatal hypothermia remains highly prevalent despite most units maintaining ward temperatures above WHO minimum standards (26°C). Strengthening all components of the warm chain, particularly at night and during colder seasons, is essential to reduce hypothermia and improve survival.

**Key Findings:** 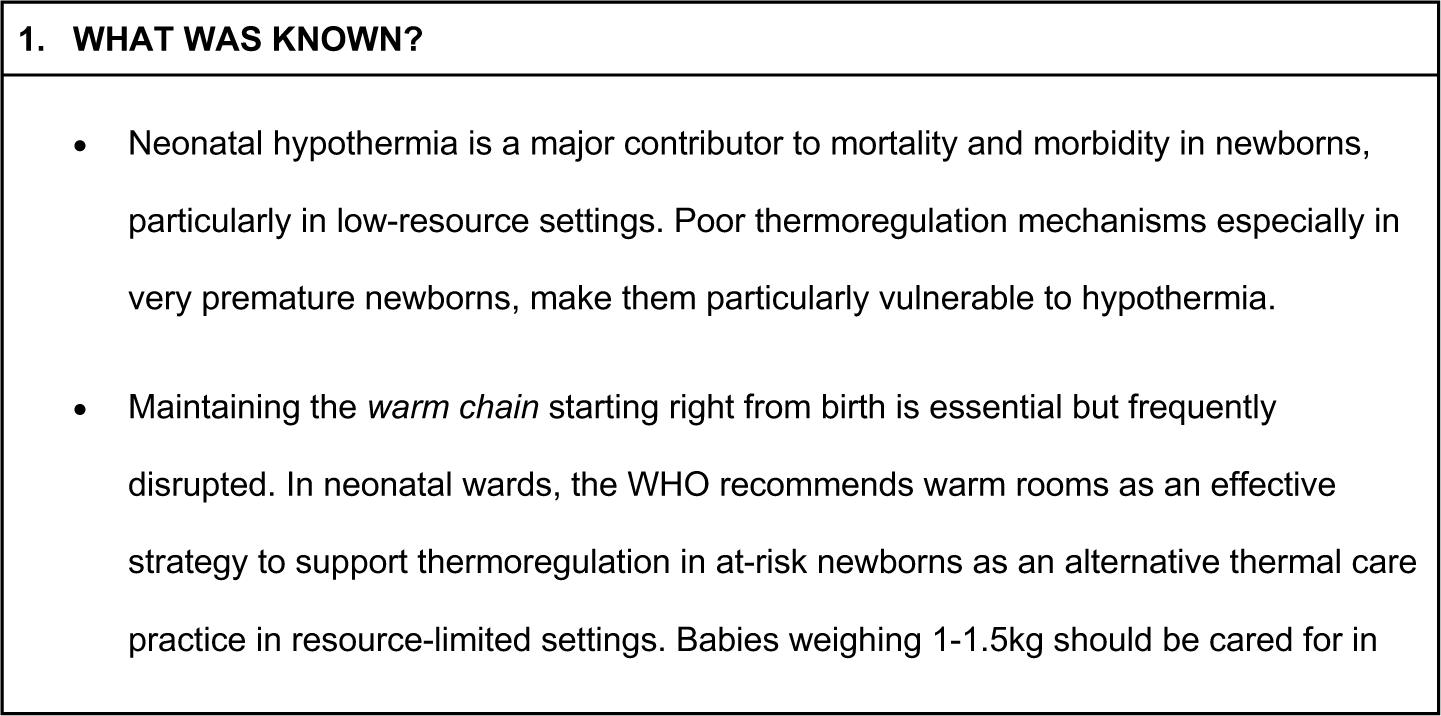

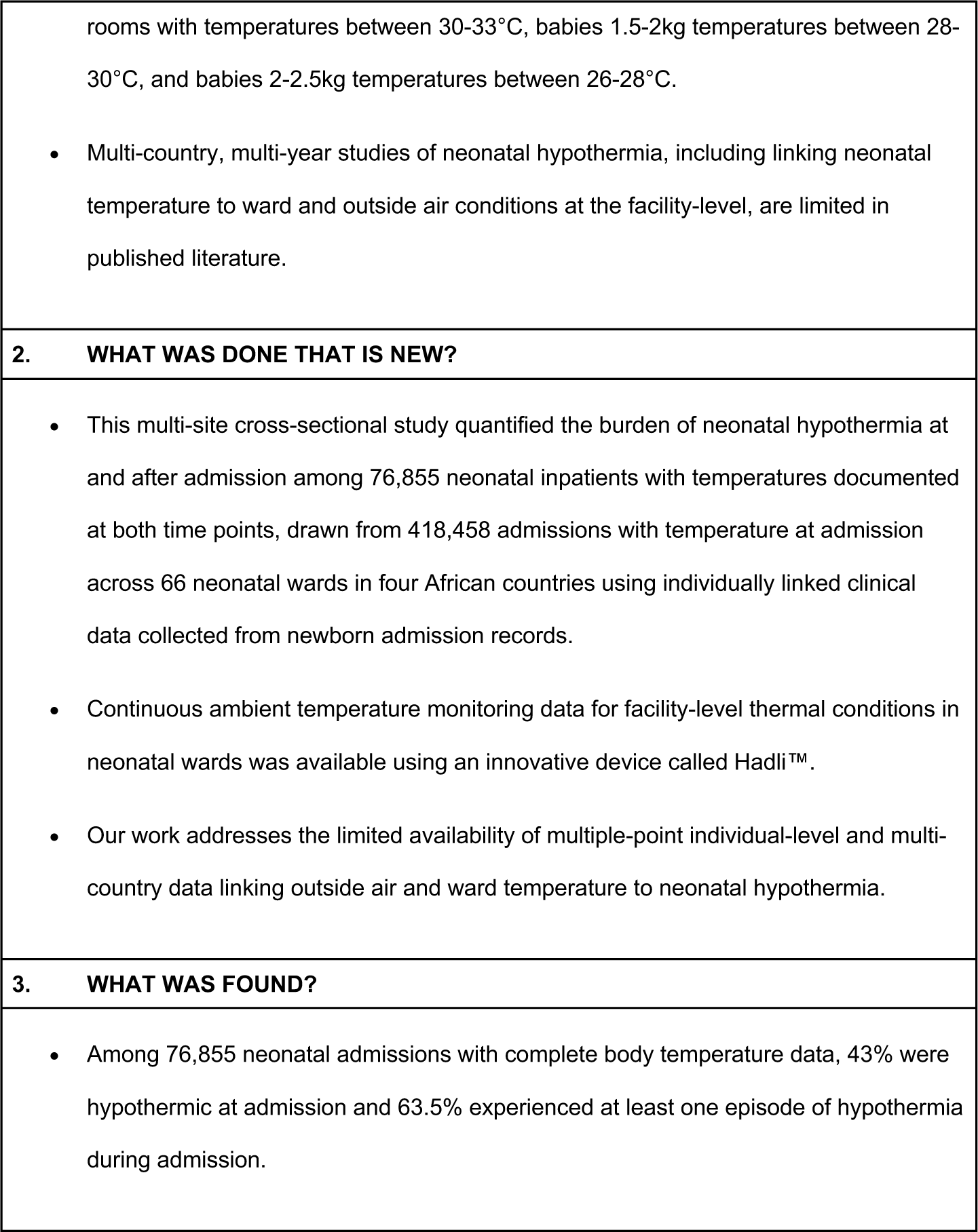

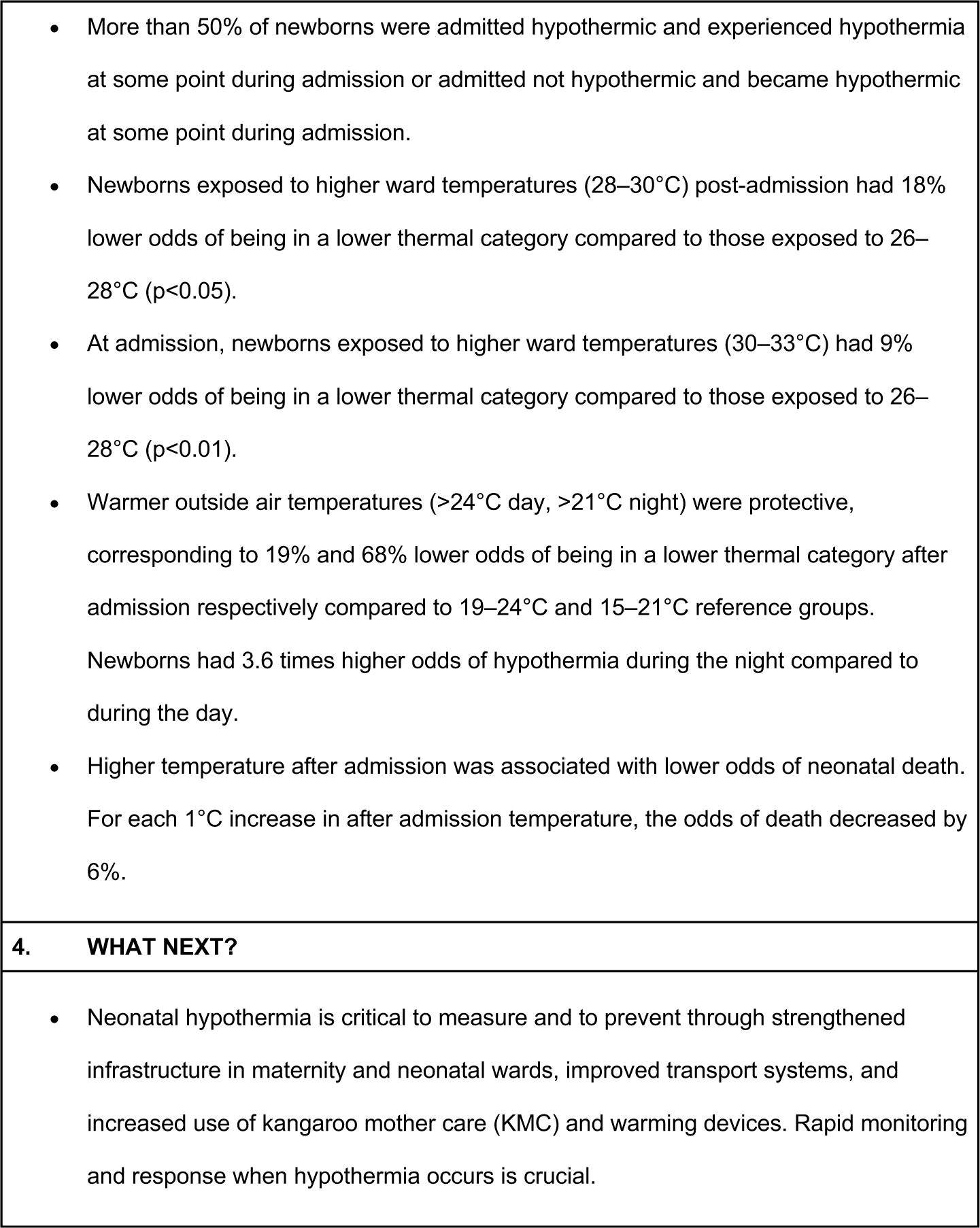

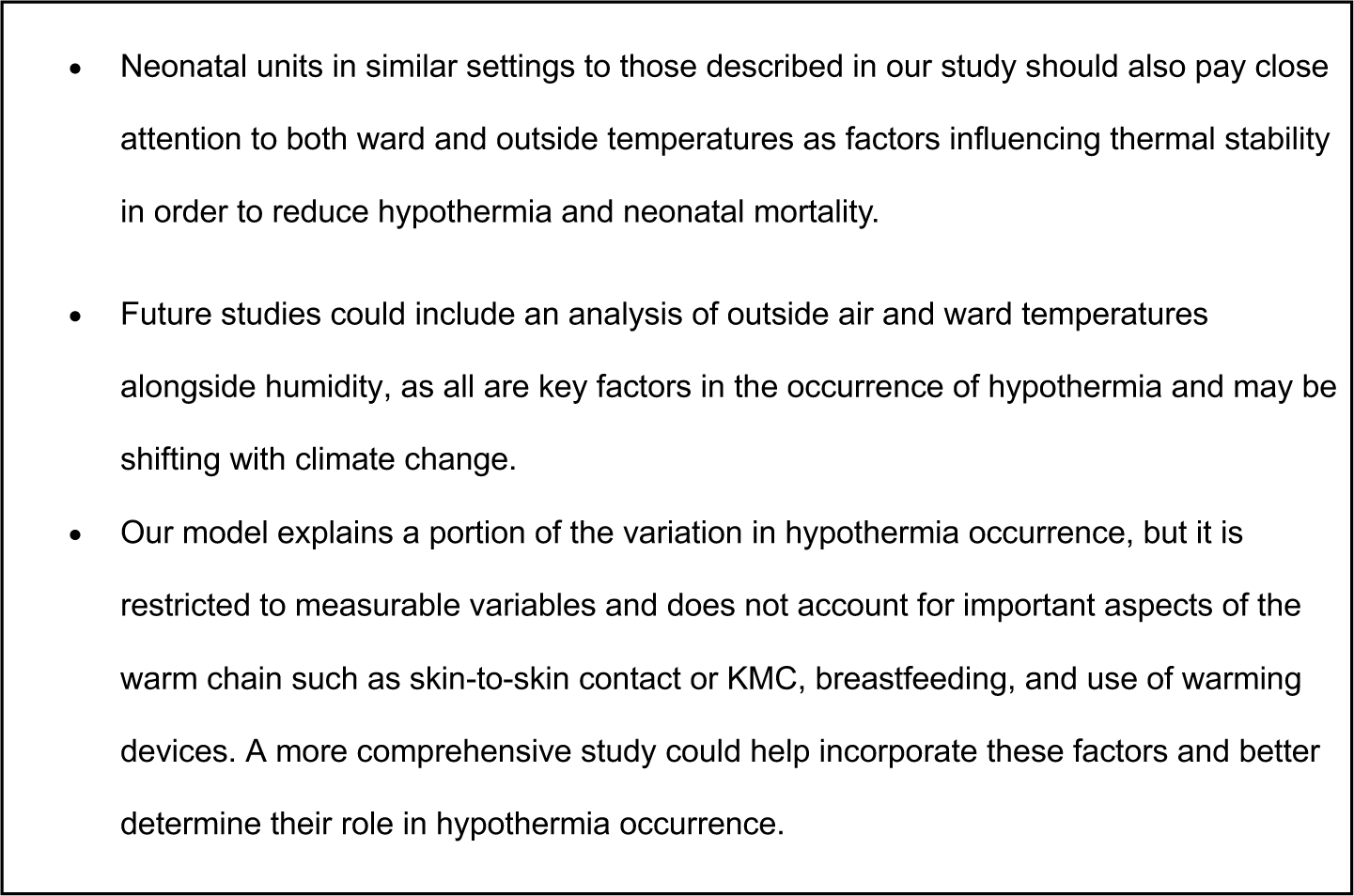

## Introduction

Every year, an estimated 2.3 million newborns die globally, representing nearly 50% of under-five deaths [1]. Over one million of these deaths occur in sub-Saharan Africa, mostly due to preterm birth complications [2,3]. Hypothermia (mild: 36.0–36.4°C, moderate: 32.0–35.9°C, severe: <32.0°C) is a major contributor to morbidity and mortality in newborns [4–6]. Poor thermoregulation mechanisms in newborns, especially in premature newborns, make them especially vulnerable to hypothermia [7].

Achieving Sustainable Development Goal (SDG) 3.2 of reducing neonatal mortality below 12 per 1000 live births by 2030 requires improved prevention of hypothermia [8]. WHO and UNICEF’s Every Woman, Every Newborn, Everywhere (EWENE, previously Every Newborn Action Plan) targets 80% of districts with at least one hospital providing inpatient newborn care including respiratory support [9]. High-quality inpatient care, including thermal management, can avert the majority of neonatal deaths [10].

Preventing hypothermia (<36.5°C) is promoted by the WHO’s ‘warm chain’ which includes a delivery room temperature ≥26°C, immediate drying, skin-to-skin contact, early breastfeeding, appropriate wrapping, use of a radiant warmer when indicated, delayed bathing and weighing, and warmth during transport. Breaks in this chain can cause rapid heat loss [5].

Neonatal hypothermia increases morbidity and mortality, with complications including respiratory distress, metabolic instability, infections, and poor feeding [11]. Affected infants may require prolonged hospital stay, extended respiratory support [12], and have poorer outcomes on Continuous Positive Airway Pressure [6]. Hypothermia has been associated with a four-fold increase in mortality, noting that the most vulnerable babies are most likely to be cold as well as most likely to die [13].

WHO recommends delivery room temperatures of 25-28°C [5] and graded warm rooms in neonatal units: infants 1-1.5kg require rooms 30-33°C, those 1.5-2kg need 28-30°C, and those 2-2.5kg need 26-28°C [5]. Inadequate outside air temperature undermines warm chain practices [15], making continuous monitoring and optimizing ward temperature a critical component of thermal care. In low-resource settings, delivery rooms and neonatal wards often operate with temperatures below 26°C due to a lack of temperature control, unreliable electricity, or architectural design. Seasonal variations, particularly colder or rainy months, can lower ward temperatures and increase hypothermia risk even in tropical climates [14]. Transfers from labor wards to neonatal wards or between hospitals further expose newborns to outside temperatures if transport is not adequately prepared [15].

This study described neonatal hypothermia at admission and during hospital stay, and its association with outside and ward temperatures in neonatal wards across four countries implementing with the NEST360 alliance. Specific objectives were to: (i) estimate prevalence and seasonal variation of neonatal hypothermia at admission and during hospital stay; (ii) describe patterns and seasonal variations in outside air and ward temperatures; and (iii) assess their associations with hypothermia across admission and inpatient stay.

## Methods

### Study Design and Study Sites

This was a retrospective observational study to examine individual-level neonatal data, and hospital-level ward and outside air temperature data from 66 neonatal units in Kenya, Malawi, Nigeria, and Tanzania. Newborn Essential Solutions and Technologies (NEST360) is a multidisciplinary alliance of 23 organizations working with Ministries of Health in Kenya, Malawi, Nigeria, Tanzania and since 2025 in Ethiopia, to reduce inpatient neonatal mortality in hospitals through provision of innovative medical devices, healthcare worker training and mentorship, and evidence-based, data-driven quality of care [16]. NEST360 support included equipping neonatal wards with radiant warmers, wall heaters, temperature monitors, and clinical training and quality improvement coaching on holistic inpatient neonatal care, including warm chain. The study is reported in accordance with the Strengthening the Reporting of Observational Studies in Epidemiology (STROBE) guidelines (see Appendix 7).

### Data Sources

Individual patient data were collected using the Newborn Inpatient Dataset (NID), which is described elsewhere [17]. We analyzed data on newborn temperature at admission from January 2021 to June 2025. The newborns’ lowest temperatures after admission were available from July 2024 to June 2025 in Tanzania, Malawi, and Nigeria; however, they were only available in Kenya after October 2024.

Ambient temperature data from neonatal wards were available through continuous Hadli™ monitors from July 2024 to June 2025 [18]. These included daily minimum, maximum, and average ward temperature readings. Additional outside air temperature data around all units between January 2021 and June 2025 were accessed from Open-Meteo [19]. These data were captured at two meters above ground at a one-hour temporal resolution and one to 11 kilometers spatial resolution.

Analyses included all available observations for each variable. Proportions were calculated using the number of neonates with non-missing data for the variable of interest as the denominator, with denominators reported alongside each result.

### Prevalence and seasonal variations of neonatal hypothermia

Newborn temperature at admission and lowest temperature after admission were used to determine each newborn’s hypothermia status at admission and during their hospital stay, respectively.

To evaluate transitions in hypothermia status from admission to hospital stay, we grouped all newborn records into four categories based on hypothermia statuses at both points in time: those admitted not hypothermic who remained not hypothermic, those admitted not hypothermic who became hypothermic at some point, those admitted hypothermic who also experienced hypothermia at some point during admission, and those admitted hypothermic followed by normothermia during admission.

### Patterns and seasonal variations in outside air and ward temperatures

Descriptive analyses were conducted to identify patterns in outside air temperatures across multiple years and by country. Further analyses examined isolated seasonal effects within each country. Hourly outside air readings were stratified into day and night periods, and then summarized into minimum, average, and maximum readings.

A correlation analysis was used to determine the relationship between outside air temperatures and neonatal ward temperatures.

### Associations between outside air and ward temperatures with neonatal hypothermia

Correlation analyses were used to determine the relationship between both outside air and ward temperatures with neonatal hypothermia, at admission and during hospital stay. Since lowest body temperatures recorded per baby did not have a corresponding date, outside air temperatures and minimum ward temperatures were aggregated over length of hospital stay in days for each baby following the newborn’s date of admission to the neonatal ward. Minimum temperatures were used because they represent the most critical thermal conditions in which newborns are most susceptible to hypothermia.

Multivariate ordinal logistic regression was used to assess associations between outside air temperatures, ward temperatures, and hypothermia at admission and during hospital stay, adjusting for birth weight and birth location when appropriate. The outcome variable was defined as a three-level ordinal measure of hypothermia severity: no hypothermia (normal), mild hypothermia, and moderate-to-severe hypothermia. Seasonality was included in the model using sine and cosine terms to capture cyclical patterns in newborn hypothermia.

A severity score, adapted from validated clinical severity scores for neonatal mortality was developed using three clinical indicators at admission: gestational age, oxygen saturation, and body temperature [20, 21, 22]. Each variable was grouped into categories representing escalating levels of risk and assigned a score from 1 to 3, with higher scores indicating greater physiological instability. Scores across the three indicators were summed to create a composite severity score. For our models predicting hypothermia severity as the outcome, admission body temperature was excluded from the severity score. This composite score was assessed through sensitivity analyses described in the supplementary materials.

### Associations between neonatal characteristics and body temperature with neonatal mortality

Multivariate logistic regression was used to examine associations between neonatal characteristics, including primary diagnosis and lowest post-admission temperature, and neonatal mortality. The severity score described above was also included in this model using the original three components, which have been validated as important predictors of neonatal mortality.

## Results

### Overview

Between 1 January 2021 and 30 June 2025, 444,580 newborns were admitted across all 66 neonatal units included in the study. Neonatal characteristics across each country are described in Appendix 1. Between 1 July 2024 and 30 June 2025, 101,956 newborns were admitted across all 66 neonatal units with temperature recorded at admission. Of these, 76,855 (75.4%) had temperatures documented at both admission and subsequent measurement after admission. For the analysis of associations between neonatal, ward, and outside air temperatures, Hadli™ data were available from 59 units, yielding a study sample of 51,181 newborns who met the inclusion criteria.

### Prevalence and seasonal variations of neonatal hypothermia

*Hypothermic at Admission:* Among 444,580 neonatal ward admissions across 66 hospitals in Malawi, Tanzania, Nigeria, and Kenya, 94.1% (N=418,458) had temperature at admission documented. Of these, 47.3% (N=220,684) reported a hypothermic temperature (<36.5°C) at admission across the study period (51.0% in 2021 and 44.2% in 2024). The proportion of hypothermic admissions varied between 61.9% in Malawi and 22.8% in Tanzania. Although overall trends suggested sustained decreases in the proportion of hypothermic admissions, program and country-level analyses revealed seasonal fluctuations in hypothermic admission rates over the years. (Figure 1). When stratified by birthweight and birth location, there were more pronounced fluctuations in hypothermic admissions within specific subgroups, although all groups demonstrate an overall yearly decrease (Appendix 2 and 3).

**Figure 1:**
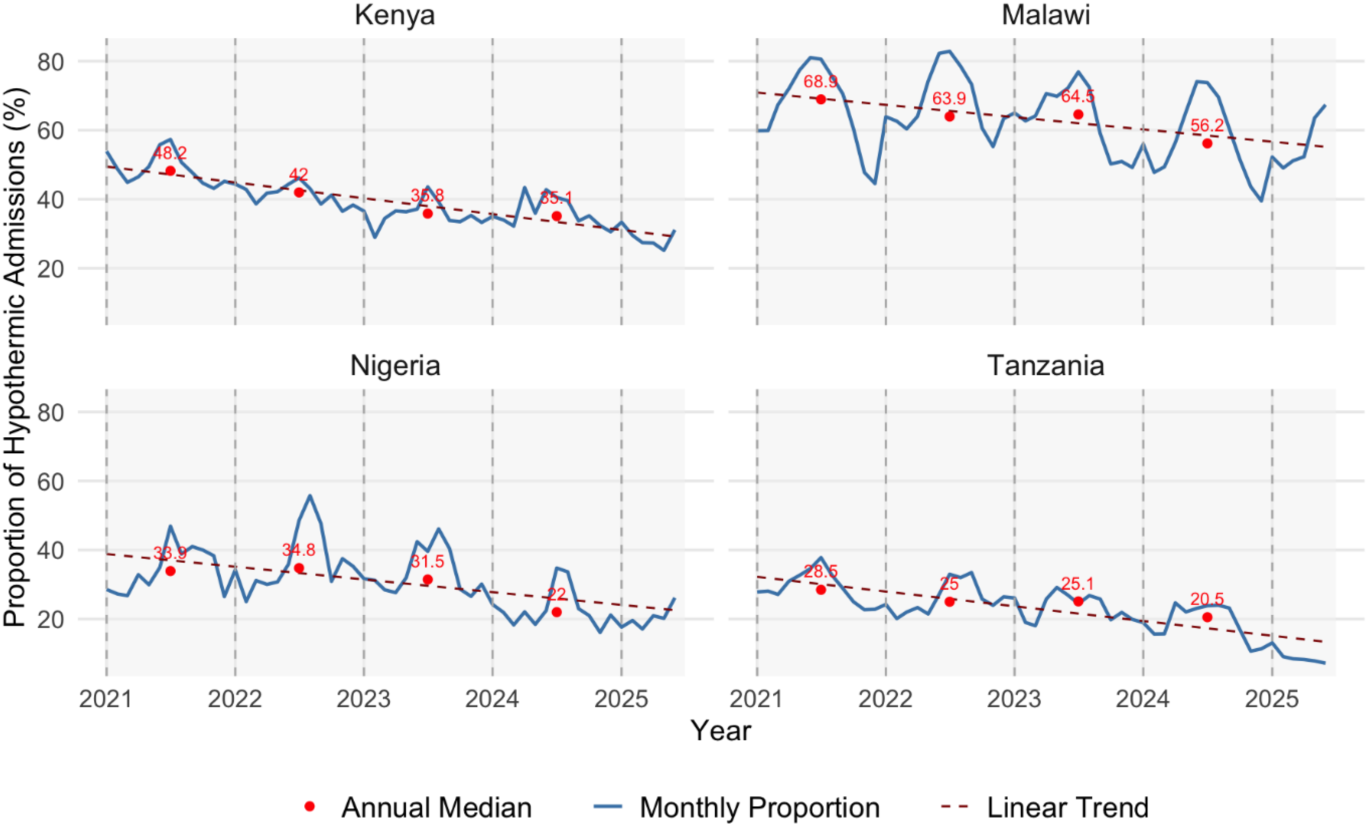
Monthly proportion of hypothermic admissions pooled at the country level showing trends January 2021–June 2025. N=418,458.

Month-to-month fluctuations for hypothermic admissions were apparent, with mid-year months (June, July, and August) consistently among the highest across years. Despite recurring seasonal peaks, hypothermia admissions declined overall, with an average annual reduction in hypothermia admissions of 2.27% (2.67%, 3.30%, 4.23%, and 4.43% in Malawi, Tanzania, Kenya, and Nigeria, respectively). The mid-year rate of increase was 2.32% (48.2% in 2021 to 59.8% in 2025).

*Hypothermic after Admission:* Among all neonatal ward admissions across 66 hospitals in four countries, 76.5% (n=77,987) had data on the lowest temperature after admission. 63.5% (n=48,746) of these records reported a hypothermic temperature (<36.5°C) after admission across the study period. The proportion of hypothermic newborns during hospital stay varied widely, from 74.4% in Malawi to 18.5% in Tanzania.

*Inborn and Outborn Hypothermia at and after Admission:* At admission, 46.1% (45.6-46.5%) of inborn neonates were hypothermic compared to 34.1% (33.5-34.8%) of outborn neonates. After admission, 63.1% (62.7-63.5%) of inborn neonates experienced hypothermia compared to 63.7% (63-64.4%) of outborn neonates. The proportion of inborn hypothermia at admission varied widely, from 59.0% in Malawi to 9.3% in Tanzania. The proportion of inborn hypothermia during the hospital stay also varied widely, from 73.7% in Malawi to 16.4% in Tanzania (Figure 2).

**Figure 2:**
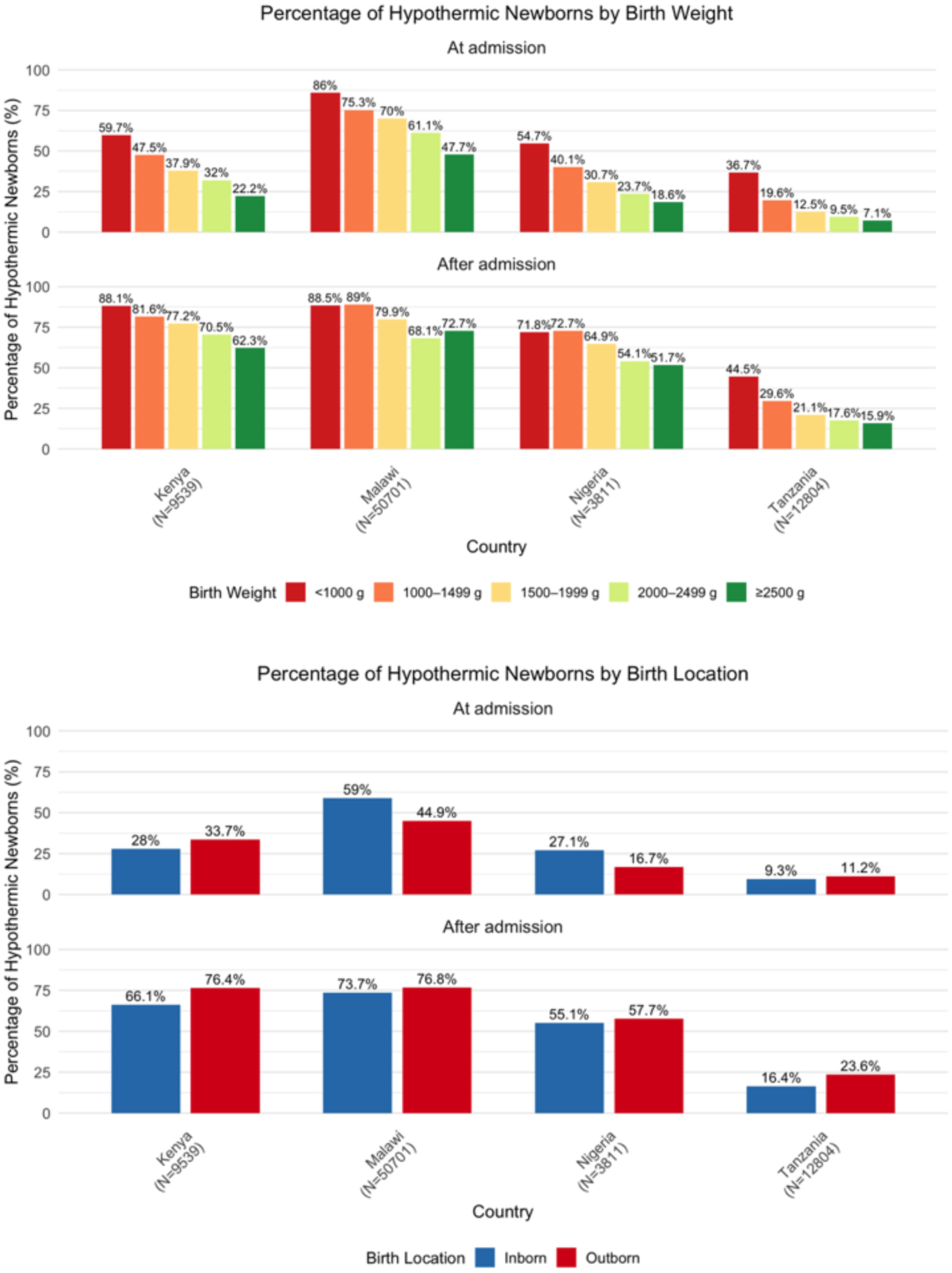
Hypothermia at and after admission by birth weight and birth location for hypothermic newborns in Malawi, Nigeria, Tanzania (1 July 2024 to 30 June 2025) and Kenya (1 November 2024 to 30 June 2025).

*Birthweight Group Hypothermia at and after Admission:* Newborns in the lowest birth weight categories (<1000g and 1000-1499g) consistently had the highest rates of hypothermia at and after admission across all countries. At admission, the proportion of newborns <1000g with hypothermia ranged from 36.7% in Tanzania to 86% in Malawi. After admission, this ranged from 44.5% in Tanzania to 88.5% in Malawi (Figure 2).

*Severity of Hypothermia at and after Admission:* Among all neonatal ward admissions, 15.8% of neonates experienced mild hypothermia, 27.0% experienced moderate hypothermia, and 0.2% experienced severe hypothermia at admission. The proportion of moderate hypothermia at admission varied widely, ranging from 37.6% in Malawi to 3.2% in Tanzania. After admission, about 34.9% of neonates experienced mild hypothermia, 28.4% experienced moderate, and 0.1% experienced severe hypothermia. The proportion of moderate hypothermia during the hospital stay also varied widely, from 35.8% in Malawi to 3.7% in Tanzania (Figure 3, Appendix 4).

**Figure 3:**
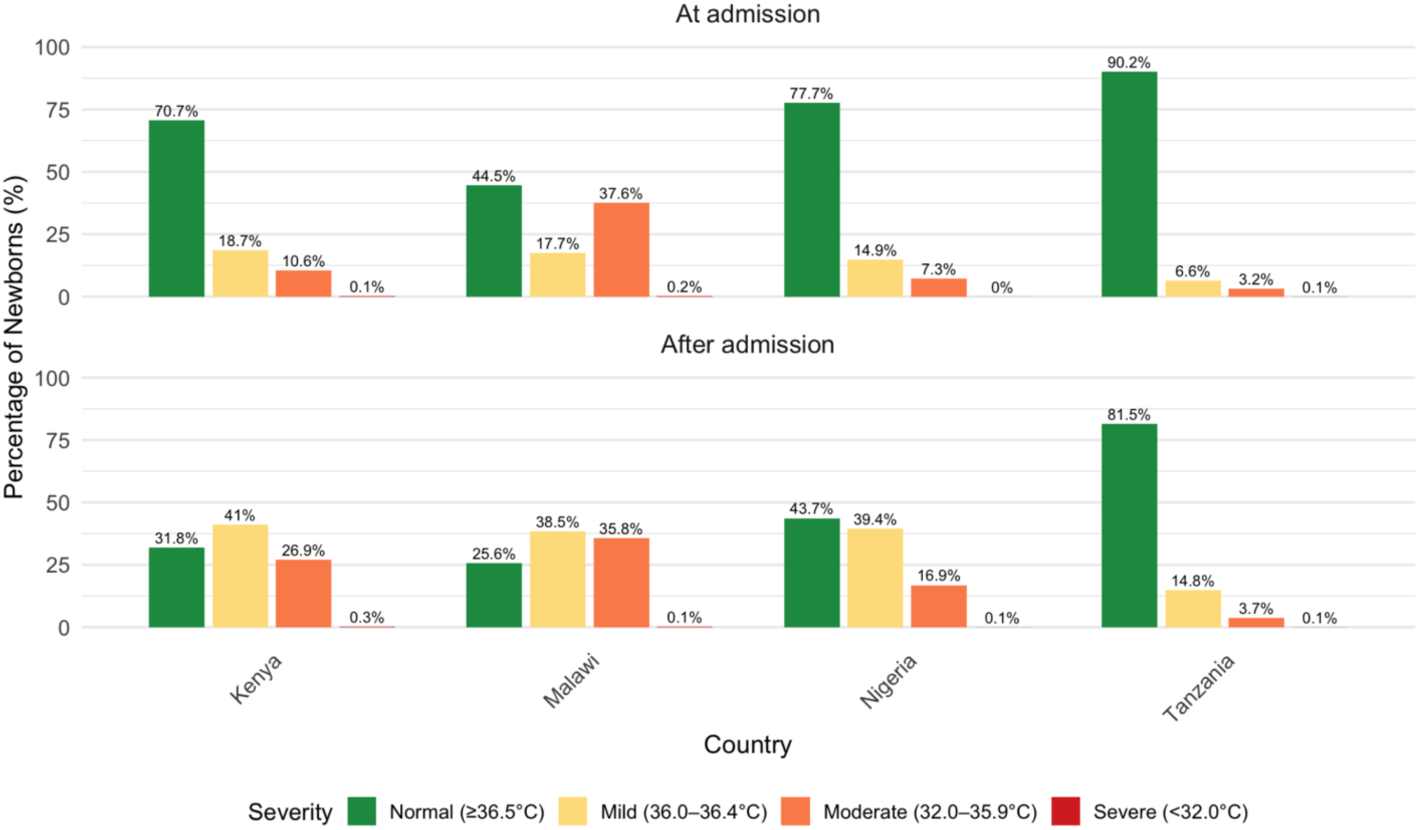
Severity of hypothermia at and after admission for hypothermic newborns in Malawi, Nigeria, Tanzania (1 July 2024 to 30 June 2025) and Kenya (1 November 2024 to. 30 June 2025).

*Transitions in Hypothermia Status:* More than 50% of newborns were admitted hypothermic and experienced hypothermia at some point during admission or admitted not hypothermic and became hypothermic at some point during admission. Among all neonatal ward admissions, 75.4% (n=76,855) had both temperature at and after admission documented. Forty three percent (n=33,021) of these records reported a hypothermic temperature (<36.5°C) at admission across the study period (55.5% in Malawi and 9.8% in Tanzania). After admission, 63.5% (n=48,746) reported a hypothermic lowest temperature across the study period (74.4% in Malawi and 18.5% in Tanzania). Only 27.9% of newborns were not hypothermic at admission and remained so during inpatient stay, while 8.7% were hypothermic at admission and became not hypothermic. Almost a third (29.2%), were admitted not hypothermic but became hypothermic, and 34.3% were admitted hypothermic and remained so. Very few (country range: 3.5%-11.3%) were admitted hypothermic and transitioned to not hypothermic (Table 1).

**Table 1:** Neonatal thermal status transitions at and after admission for Malawi, Nigeria, Tanzania (1 July 2024 to 30 June 2025) and Kenya (1 November 2024 to 30 June 2025).

|  | Program Level<br>N (%) | Kenya<br>N (%) | Malawi<br>N (%) | Nigeria<br>N (%) | Tanzania<br>N (%) |
| --- | --- | --- | --- | --- | --- |
|  | N=76,855 | N=9,539 | N=50,701 | N=3,811 | N=12,804 |
| Hypothermic at admission | 33,021 (43) | 2,797 (29.3) | 28,122 (55.5) | 848 (22.3) | 1254 (9.8) |
| Hypothermic after admission | 48,746 (63.5) | 6,505 (68.2) | 37,724 (74.4) | 2,146 (56.3) | 2,371 (18.5) |
| Admitted not hypothermic, did not experience hypothermia during admission | 21,422 (27.9) | 2,699 (28.3) | 7,228 (14.3) | 1,516 (39.8) | 9,979 (77.9) |
| Admitted not hypothermic, experienced hypothermia during admission | 22,412 (29.2) | 4,043 (42.4) | 15,351 (30.3) | 1,447 (38.0) | 1,571 (12.3) |
| Admitted hypothermic, did not experience hypothermia during admission | 6,687 (8.7) | 335 (3.5) | 5,749 (11.3) | 149 (3.9) | 454 (3.5) |
| Admitted hypothermic, experienced hypothermia during admission | 26,334 (34.3) | 2,462 (25.8) | 22,373 (44.1) | 699 (18.3) | 800 (6.2) |

Seasonal trends were observed over the most recent year, with higher proportions of newborns who were admitted not hypothermic and became hypothermic or admitted hypothermic and experienced hypothermia during admission during mid-year months (June to September), followed by a decline in the final months of the year and a subsequent rise (Figure 4). In contrast, newborns who were admitted not hypothermic and remained not hypothermic or admitted hypothermic and became not hypothermic maintained relatively stable proportions throughout the year (Figure 4). Across countries and months of admission, notable differences were observed (Appendix 5).

**Figure 4:**
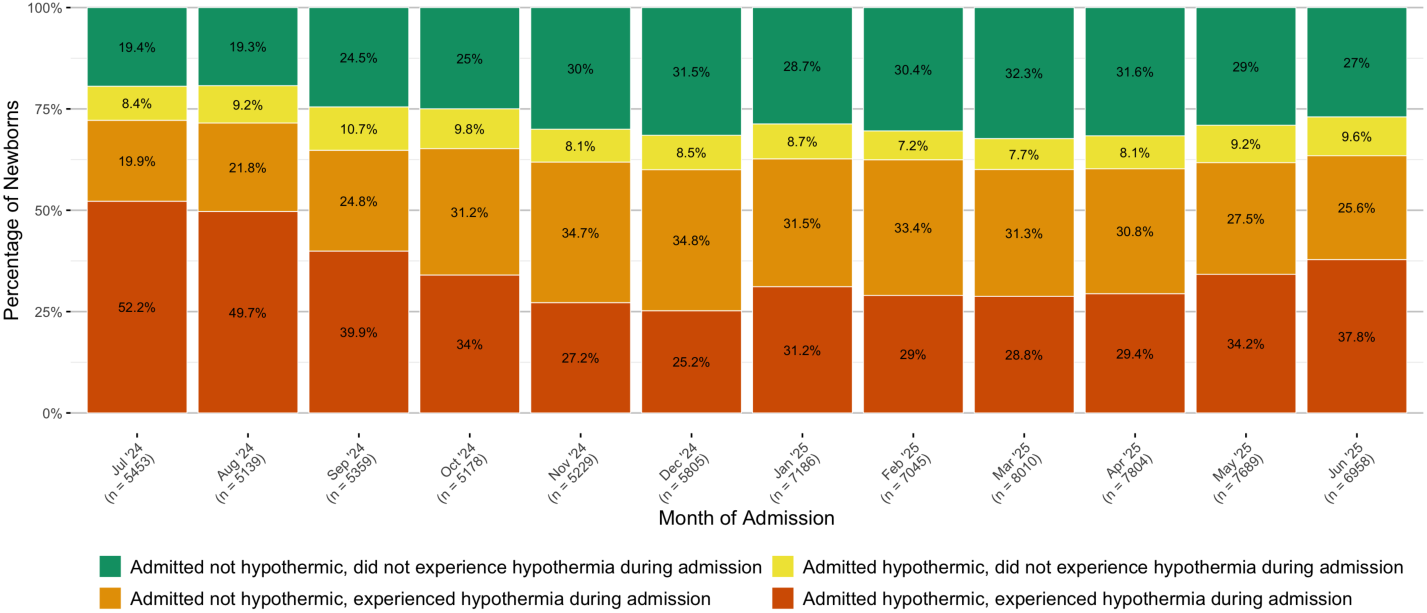
Neonatal thermal status transitions across 12 months of admission for Malawi, Nigeria, Tanzania (1 July 2024 to 30 June 2025) and Kenya (1 November 2024 to 30 June 2025). N = 76,855.

### Patterns and seasonal variations in outside air and ward temperatures

Mean day and night outside air temperatures varied by country: 24.7 °C (SD 3.9) (22.3°C in Kenya to 28.7°C in Nigeria) and 19.7 °C (SD 4.1) (16.4°C in Kenya to 24.9°C in Nigeria), respectively. The average difference between temperatures was 5.1°C (SD = 1.9°C) (3.7°C in Nigeria to 5.9°C in Kenya). At the country level, seasonal variations in day and night outside air temperatures were evident (Figure 5).

**Figure 5:**
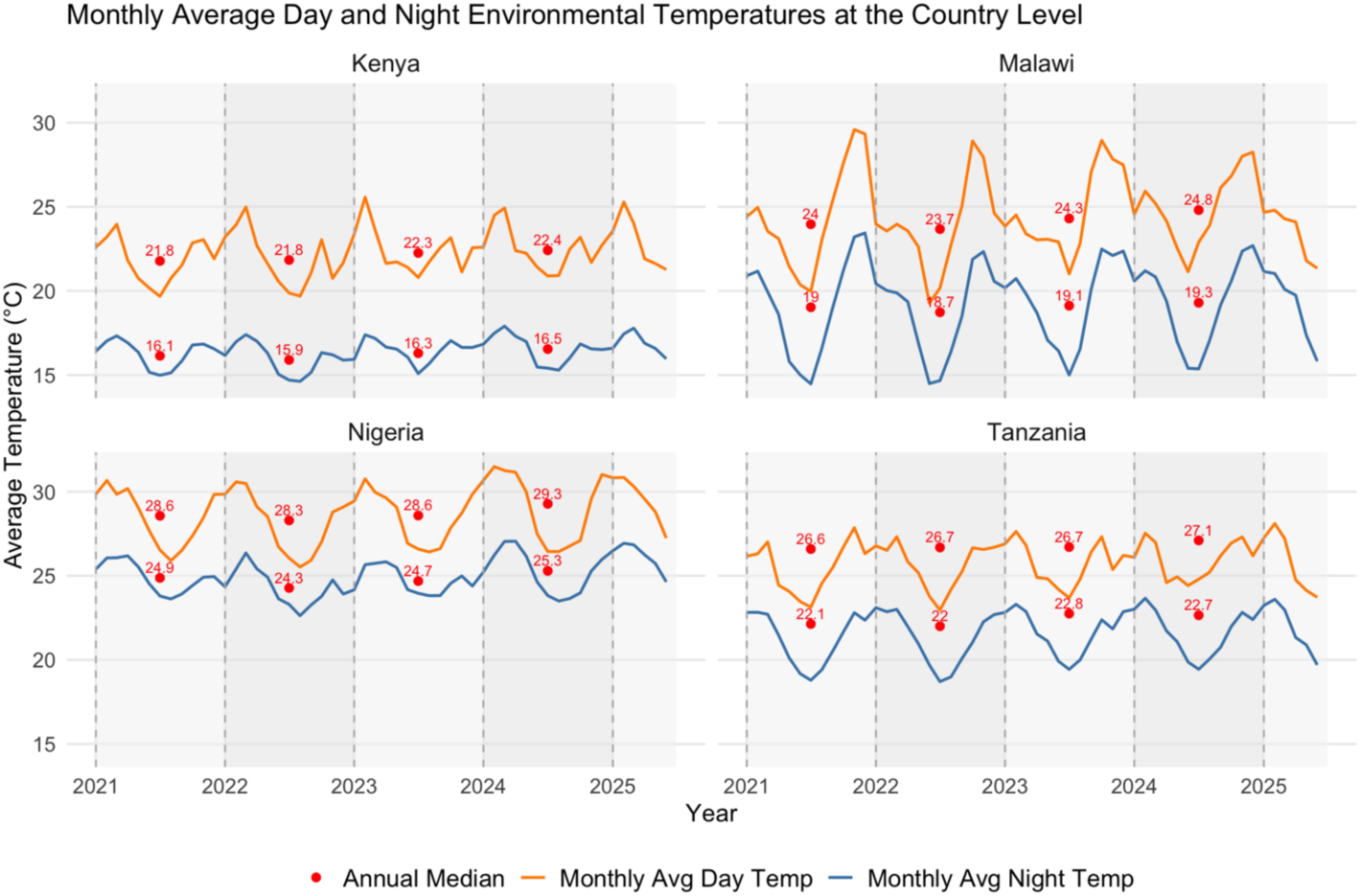
Monthly average day and night outside air temperatures at the country level across years (January 2021 to June 2025). N = 2,325,072.

Overall, average, minimum and maximum ward temperatures were 30.6 °C (SD = 2.1°C), 29.3°C (SD = 2.1°C) and 32.1°C (SD = 2.3°C), respectively. The average difference between maximum and minimum temperature was 2.8°C (SD = 1.5°C), lowest in Nigeria (2.1°C) and highest in Malawi (3.2°C) (Appendix 6). Across countries, average maximum temperatures ranged from 31.0°C (Nigeria) to 32.5°C (Malawi) and average minimum temperatures ranged from 28.9°C (Nigeria) to 29.6°C (Kenya) (Appendix 6). At the country level, seasonal variations in neonatal ward temperatures were evident (Figure 6).

**Figure 6:**
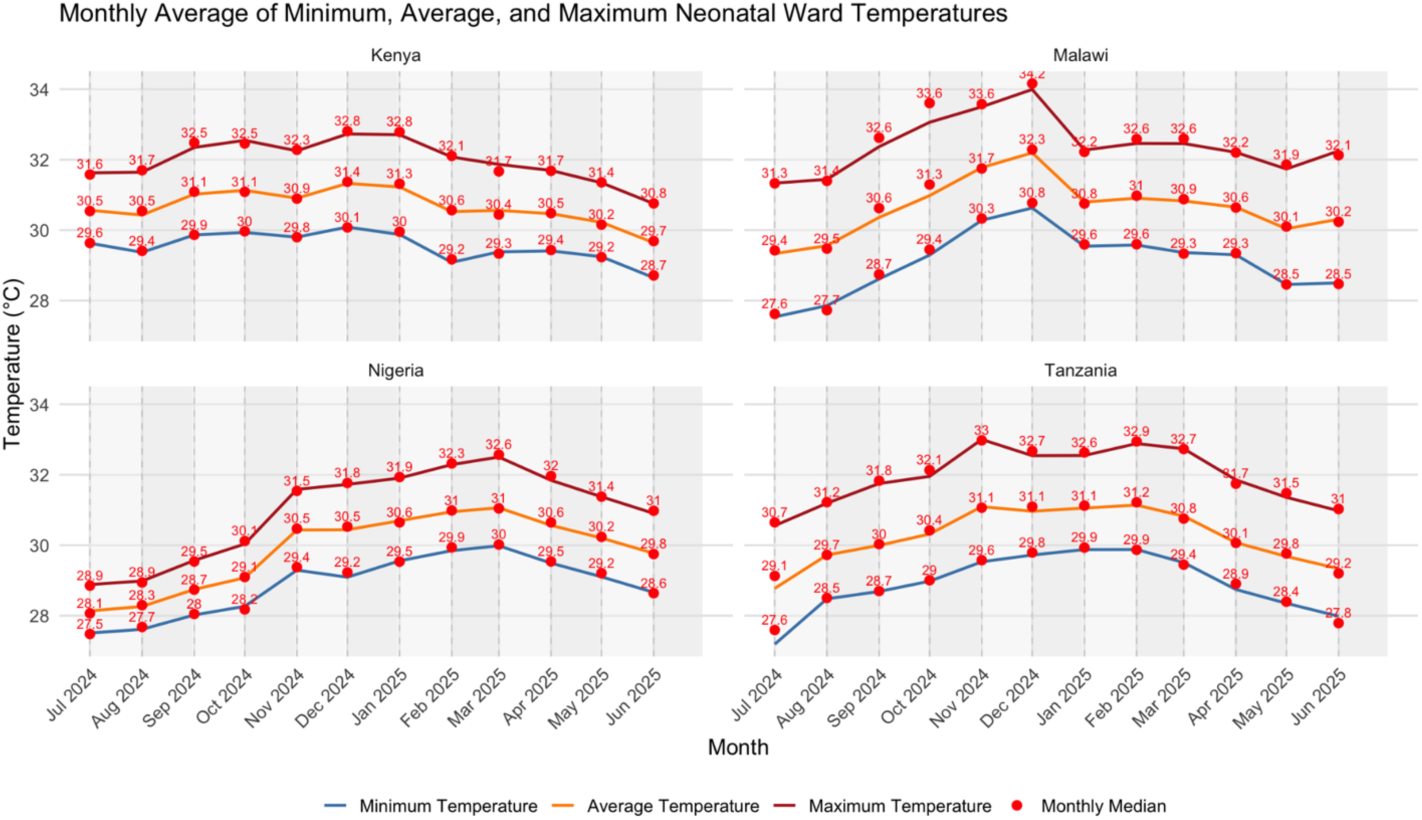
Monthly average of minimum, average, and maximum neonatal ward temperatures at the country level (1 July 2024 to 30 June 2025). N= 16,212.

Pearson’s correlation showed that average ward temperatures in the neonatal wards were significantly correlated with both average day (r = 0.18, p < 0.00001) and night (r = 0.14, p < 0.00001) outside air temperatures.

### Associations between outside air and ward temperatures with neonatal hypothermia

Pearson’s correlation showed highly significant but weak positive associations between newborn temperatures at admission and average outside air day and night temperatures, and the same pattern was observed for the lowest temperature after admission. The lowest temperature after admission demonstrated positive but weak associations with the average, minimum, and maximum ward temperatures during hospital stay (Table 2).

**Table 2:** Correlations between outside air, ward, and newborn temperatures at and after admission. \**Temperatures calculated over the actual length of hospital stay*.

| Source | Measure Timing | Variable Type | Correlation |
| --- | --- | --- | --- |
| Outside Air | Admission Temp | Day Avg | 0.25 |
|  |  | Night Avg | 0.25 |
|  | Lowest Temp after Admission | Day Avg* | 0.20 |
|  |  | Night Avg* | 0.25 |
| Ward | Admission Temp | Avg | 0.13 |
|  |  | Min | 0.14 |
|  |  | Max | 0.11 |
|  | Lowest Temp after Admission | Avg* | 0.12 |
|  |  | Min* | 0.11 |
|  |  | Max* | 0.12 |

*Hypothermia severity ordinal logistic regression modeling:* At admission, newborns exposed to higher ward temperatures during ward stay (30-33°C) had 9% lower odds of being in a lower thermal category (Normal, Mild Hypothermia, Moderate/Severe Hypothermia), compared with those exposed to 26-28°C (OR: 0.91, p<0.01). Exposure to lower (<26°C) and moderately higher (28-30°C) temperatures did not substantially differ from the reference category. This suggests that the highest temperature range (30-33°C) is associated with the greatest protective effect at admission. Warmer outside air temperatures (>24°C and >21°C for day and night respectively) corresponded to 40% and 61% lower odds of being in a lower thermal category at admission in comparison to 19–24°C and 15–21°C.

After admission, newborns exposed to higher ward temperatures during ward stay (28-30°C and 30-33°C) had 18% (p<0.05) and 15% (approaches statistical significance; p = 0.06) lower odds respectively of being in a lower thermal category, compared with those exposed to 26-28°C (Table 3). The protective effect appears to diminish at higher temperatures, with a smaller reduction in odds observed in the 30-33°C category compared to 28-30°C. Warmer outside air temperatures (>24°C and >21°C for day and night respectively) corresponded to 19% and 68% lower odds of being in a lower thermal category after admission in comparison to the same reference groups mentioned above (Table 3).

**Table 3:** Ordinal logistic regression model for newborn hypothermia after admission. Adjusted OR with 95% CIs shown. *p-value <0.05, **p-value <0.001, ***p-value <0.00001, all others without statistical evidence of a relationship.

| Variable | Adjusted Odds Ratio and CI<br>N = 51,181 \| R<sup>2</sup> = 0.32 |
| --- | --- |
| Neonatal Temperature at Admission |  |
|  | 0.76 (0.75–0.77)*** |
| Ward Temperatures (Hospital Stay) \| 26–28°C (reference) |  |
| Ward min temp <26°C | No Observations |
| Ward min temp 28–30°C | 0.82 (0.70–0.96)* |
| Ward min temp 30–33°C | 0.85 (0.71–1.01) |
| Outside Air Temperatures (Day) \| 19–24°C (reference) |  |
| Daytime min temp ≤19°C | 1.00 (0.95–1.05) |
| Daytime min temp >24°C | 0.81 (0.75–0.86)*** |
| Outside Air Temperatures (Night) \| 15–21°C (reference) |  |
| Nighttime min temp ≤15°C | 1.11 (1.05–1.17)*** |
| Nighttime min temp >21°C | 0.32 (0.30–0.34)*** |
| Birth Weight Categories \| 2500–4000 g (reference) |  |
| <1000 g | 2.38 (2.04–2.78)*** |
| 1000–1499 g | 2.15 (1.97–2.35)*** |
| 1500–1999 g | 1.30 (1.23–1.37)*** |
| 2000–2499 g | 0.87 (0.83–0.91)*** |
| >4000 g | 0.51 (0.45–0.58)*** |
| Severity Score |  |
|  | 1.08 (1.06–1.11)*** |
| Seasonal Effects |  |
| Sine component of seasonal effect | 0.87 (0.85–0.89)*** |
| Cosine component of seasonal effect | 1.15 (1.11–1.18)*** |

**Table 4:** Logistic regression model for newborn discharge outcome. Adjusted OR with 95% CIs shown. *p-value <0.05, **p-value <0.001, ***p-value <0.00001, all others without statistical evidence of a relationship.

| Variable | Adjusted Odds Ratio and CI<br>N = 60,232 \| R<sup>2</sup> = 0.20 |
| --- | --- |
| Respiratory Distress Syndrome |  |
|  | 1.33 (1.26–1.40)*** |
| Outborn Status |  |
|  | 1.82 (1.69–1.96)*** |
| Birth Weight Category \| Reference: 2500–4000 g |  |
| <1000 g | 31.52 (26.13–38.02)*** |
| 1000–1499 g | 7.52 (6.62–8.55)*** |
| 1500–1999 g | 2.21 (1.96–2.50)*** |
| 2000–2499 g | 1.16 (1.04–1.29)** |
| >4000 g | 0.71 (0.52–0.97)* |
| Primary Diagnosis Category \| Reference: Prematurity |  |
| Congenital Malformations | 4.03 (3.43–4.74)*** |
| Infection | 1.56 (1.36–1.78)*** |
| Intrapartum-related | 2.85 (2.54–3.20)*** |
| Jaundice (Pathological) | 0.43 (0.32–0.58)*** |
| Other (None of Above) | 0.80 (0.68–0.93)** |
| Lowest Recorded Temperature (°C) |  |
|  | 0.94 (0.91–0.98)** |
| Severity Score |  |
|  | 1.60 (1.55–1.65)*** |

*Discharge outcome logistic regression modeling:* Higher temperature after admission was associated with lower odds of neonatal death. For each 1°C increase in after admission temperature, the odds of death decreased by 6%. Newborns diagnosed with respiratory distress syndrome had higher odds of death compared with those without RDS. Similarly, outborn newborns had substantially higher odds of death compared with inborn newborns. The severity score, which reflects lower oxygen saturation and body temperature at admission as well as gestational age, was independently associated with increased mortality, with each unit increase corresponding to 60% higher odds of death.

## Discussion

This multi-country analysis reveals that neonatal hypothermia is an important and persistent issue in neonatal wards across Kenya, Malawi, Nigeria, and Tanzania where the majority of newborns are hypothermic upon admission to the hospital or during their ward stay. Our findings demonstrate how outside air temperatures and ward temperatures can influence neonatal temperature and hypothermia outcomes, both at admission and during hospital stay. Higher outside air temperatures and ward temperatures alike were found to reduce the occurrence of newborn hypothermia at and after admission.

Over the study period, the proportion of newborns who were hypothermic at admission steadily decreased across all countries, thus demonstrating the success of ongoing interventions to improve thermal care practices [16]. Kenya showed the greatest improvement in reducing hypothermic admissions over time, but Tanzania achieved the highest overall reduction albeit improvements were less pronounced, presumably due to lower baseline levels of hypothermia at admission. Nigeria also reached relatively high reductions, while Malawi continues to show more limited progress. Seasonal fluctuations in hypothermic admissions were observed across all countries, which emphasizes the need for seasonally adapted thermal care practices such as increased kangaroo mother care and more frequent temperature monitoring during colder periods. In each case, reductions were highest during mid-year months, likely reflecting seasonal climatic shifts. A study in Nepal found that, across all study years, newborns were less likely to exhibit axillary temperatures below 36.0°C during the months of July and August, whereas colder temperatures in January were associated with a higher prevalence of hypothermia [14].

At admission, rates of hypothermia among inborn and outborn newborns showed no clear distinction in hypothermia rates. In Nigeria and Malawi, inborn babies had higher rates of hypothermia at admission, whereas the opposite was observed in Tanzania and Kenya. However, a difference emerged in post-admission hypothermia rates. In all countries, a greater proportion of outborn than inborn babies were hypothermic. This could be linked to differences in the care provided to outborn babies upon admission to the hospital. It may reflect efforts to prevent hypothermia in outborn babies at the time of admission, but not the consistent use of thermal care practices needed to maintain normothermia throughout their hospital stay. Similar findings have been reported in Mozambique, where the importance of prolonged postnatal efforts was emphasized, as one in three neonates was hypothermic on day one regardless of admission temperature [13,15,23].

Across all countries, cases of severe hypothermia were rare, with most classified as mild or moderate. Among newborns who experienced hypothermia, the majority either experienced hypothermia both at and during admission or became hypothermic at some point during their hospital stay, which indicates that many cases develop after admission. This is likely due to gaps in sustained thermal care practices. Continued care is therefore essential to prevent the development or persistence of hypothermia [13,15,23].

Our analysis of ward temperatures revealed notable differences between daily minimum and maximum temperatures on the neonatal ward, which were particularly pronounced in some countries such as Malawi and Tanzania. Associations between ward and outside air temperatures suggest that these temperature fluctuations are partly driven by outside air conditions. Other contributing factors likely include differences in access to climate control and electricity [24,25]. Measures such as ensuring windows and doors are properly closed, well maintained, and repaired when damaged can help mitigate these temperature fluctuations.

Associations between outside air and newborn body temperatures confirmed seasonal and daily trends in hypothermia, as the relationship between newborn and outside air temperatures was stronger during colder months and more pronounced at night, consistent with previous findings [14]. Higher daytime temperatures were associated with significantly lower odds of greater hypothermia severity, whereas lower daytime temperatures had minimal effect. Conversely, higher nighttime temperatures reduced the odds of hypothermia, while lower nighttime temperatures substantially increased them. Newborn temperatures appear to be more strongly influenced by nighttime conditions, likely due to cooler external temperatures and reduced monitoring [26].

Higher ward temperatures were also associated with reduced odds of more severe hypothermia after admission. Temperatures exceeding the WHO recommended range in the ward demonstrated a protective effect for babies 2.0–2.5kg (26–28°C) against hypothermia compared to those within or below range. The WHO provides different temperature recommendations based on birth conditions in graded warm rooms: newborns weighing 1–1.5 kg require room temperatures of 30–33 °C, those 1.5–2 kg need 28–30 °C, and those 2–2.5 kg require 26–28 °C. In some cases, exceeding the recommended range may be an appropriate method to achieve adequate warmth, especially for smaller babies. The WHO describes a method known as the ‘warm room’ [5]. Given that nearly half of admitted newborns present with some degree of hypothermia, maintaining a consistently warm environment may be an effective strategy to prevent further heat loss. While maintaining warmth is essential for most newborns to prevent and reduce hypothermia, it is important to note that for those with hypoxic ischemic encephalopathy (HIE), exposure to higher temperatures can be harmful, as HIE often leads to marked thermal instability.

Extremely low birthweight infants (<1000g) exhibited 2.47 times the odds of being in a worse hypothermia category compared to infants weighing 2500-4000g. Very low birth weight infants (1000-1499g) also showed elevated risk, with more than double the odds of worse hypothermia severity. Infants weighing 1500-1999g had a more modest but still significant increase in risk, with 30% higher odds of worse thermal status at admission. In our outcome model, infants weighing <1000g also had approximately 50% higher odds of death. These findings suggest that hypothermia is both highly prevalent in this group and contributes to elevated mortality risk.

Newborns weighing less than 2000g have higher odds of hypothermia compared to those in the 2500-4000g reference group. However, infants in the 2000-2499g category show slightly lower odds relative to the reference group. This could reflect the smaller number of newborns in this category within our dataset. Another possible explanation is that moderately low birth weight infants (2000-2499g), who are recognized as at risk, may receive more proactive thermal care [27].

Higher newborn temperatures at and after admission were associated with reduced odds of mortality. Our severity score, which incorporates admission temperature, was strongly associated with mortality, with each unit increase corresponding to a 60% increase in the odds of death. Higher newborn temperatures after admission also showed a protective effect, with a 6% reduction. An observational study conducted in Mozambique discussed the relationship between admission temperature and mortality and concluded that lower admission temperatures were associated with substantially higher mortality [28].

### Strengths and Limitations

Strengths of our study include the use of standardized data collection tools (the NID, Hadli™, Context Tracker, and OpenMeteo) across all neonatal wards. The NEST360 NID dataset provided a large quantity of newborn records, available across multiple countries and numerous neonatal wards, which allowed for a robust analysis of newborn admissions. Generalizability of our findings is possible in Malawi as there is national implementation of NEST360 in all neonatal wards. Another strength is Hadli™ temperature monitoring, as it provides standard and regular temperature readings across a large number of neonatal wards. As a result, our analysis of Hadli™ data was novel and only minimally affected by missing data. Additionally, across all data sources, there is relatively little missing data.

Limitations include that Hadli™ monitors are only installed in a subset of neonatal wards and these neonatal wards had only one Hadli™ monitor, though most wards have multiple rooms. Thus, in some cases, the monitor only captures temperatures that reflect conditions experienced by some newborns. The temperature readings could also be impacted by the location of the monitor in the room. Observations in Malawi may have introduced bias in our pooled analyses as the number of observations is larger in comparison to other countries. Another potential limitation is reverse causality, as the sickest newborns are more likely to become hypothermic and to die, which could partly contribute to the observed associations.

### What is next in research

Future studies should include an analysis of outside air and ward temperatures alongside humidity (also collected through the Hadli™ monitor), as all are key factors in the occurrence of hypothermia. Furthermore, after admission neonatal temperatures are only available for the most recent year. It would be crucial to track these over a longer period of time to gain a more accurate picture of the prevalence and burden of hypothermia in the neonatal ward. Lastly, further research is required to understand the implications of WHO neonatal ward temperature recommendations for all babies and whether this range should be adapted, especially given infrastructure limitations and the inability to have neonates in multiple rooms in many low resource settings.

### Implications for clinical practice

Programs should strive to maintain warm neonatal wards throughout the year and place particular emphasis on measures to reduce fluctuations in temperatures in the hospital in colder seasons. Nighttime temperatures also require attention, as newborns may be at increased risk during this period. Our findings suggest that maintaining ward temperatures in the range of approximately 28–30°C after admission and admission temperatures around 30–33°C are associated with improved outcomes. This supports the importance of keeping neonatal rooms warm throughout admission. Though maintaining temperatures within the WHO recommended range is essential for adequate neonatal care, these efforts must be met with other interventions to maintain the warm chain, such as collaboration between labor and delivery and neonatal wards during transport and referral.

## List of abbreviations

ENAP: Every Newborn Action Plan
EWENE: Every Woman Every Newborn Everywhere
HIE: Hypoxic Ischemic Encephalopathy
NEST360: Newborn Essential Solutions and Technologies
NID: Neonatal Inpatient Dataset
RDS: Respiratory Distress Syndrome
SDGs: Sustainable Development Goals
UNICEF: United Nations Children’s Fund
WHO: World Health Organization

## Declarations

### Ethics approval and consent to participate

This study formed part of the complex evaluation of NEST360 which was granted ethical approval from the Institutional Review Boards of the London School of Hygiene & Tropical Medicine and Rice University. All collaborators received local ethical permissions for their data. No individual consent was required for the study as data were collected as part of routine data collection at each site.

### Consent for publication

Not applicable.

### Availability of data and materials

All collaborating partners in the NEST360 Alliance jointly developed and signed data sharing and transfer agreements. The NID tool, data dictionaries and training are available from NEST360/UNICEF Implementation Toolkit for Small and Sick Newborn Care [29].

### Competing interests

The authors have no competing interests to declare.

### Funding

This work was funded through the NEST360 Alliance with thanks to the Gates Foundation, ELMA Philanthropies, The Mohamed Bin Zayed Foundation for Humanity, the Beginnings Fund, Sall Family Foundation, private donors and under agreements with William Marsh Rice University. Previous support from the John D. and Catherine T. MacArthur Foundation is also gratefully acknowledged.

### Authors’ contributions

This analysis was done as part of ongoing work with the NEST360 Alliance and partners. MM, JW, and CAB developed the detailed research questions and overall analysis plan for this paper. Analysis was led by MM and supported by JW. MM, HM, JW, and CAB drafted the manuscript. EM, NR, EG, RRK, MO, and LRH provided valuable guidance and support on the manuscript development. All authors reviewed and helped to revise the manuscript. All authors reviewed and agreed on the final version. The authors’ views are their own, and not necessarily from any of the institutions they represent.

## Acknowledgements

We thank all the newborns and their families whose data were included in this analysis. We acknowledge the dedication of the healthcare workers and clinical teams across all study units whose commitment to quality newborn care made this research possible. We also recognize all hospital teams implementing with NEST360 and the data collection teams whose diligence underpins this work.

## Collaborative group

NEST360 Neonatal Inpatient Dataset and Data Systems Collaborative Group and Context Tracker

James H. Cross; Christine A. Bohne; Lucas Malla; Morris Ondieki Ogero; John Wainaina; Julius Thomas; Eric O. Ohuma; Samuel K. Ngwala; Joseph Misyenje; Msandeni Chiume; Josephine Shabani; Irabi Kassim; Prosper Mshana; Jacqueline Minja; Honorati Masanja; Franklin Okech; Vincent Ochieng; Olabisi O. Dosunmu; Rebecca E. Penzias; Kristina Shemwell; Maria Oden; Rebecca Richards-Kortum; Joy E. Lawn.

## Notes

### Competing Interest Statement

The authors have declared no competing interest.

### Author Declarations

Institutional Review Boards of the London School of Hygiene & Tropical Medicine and Rice University gave ethical approval for this work

