## Supplemental Files for "Neonatal Hypothermia at and after Admission: Burden and Associations with Outside Air Temperature and Neonatal Ward Temperature in Four Sub-Saharan African Countries Implementing with the NEST360 Alliance"

### Additional files

| | Kenya N (%) | Malawi N (%) | Nigeria N (%) | Tanzania N (%) | Total N (%) |
|---|---|---|---|---|---|
| Number of Units | N=13 | N=37 | N=9 | N=7 | N=66 |
| | N=111,224 | N=227,519 | N=19,723 | N=86,114 | N=444,580 |
| Birth weight | | | | | |
| Extremely low birth weight <1000g | 4,579 (4.1%) | 3,652 (1.6%) | 672 (3.4%) | 2,197 (2.6%) | 11,100 (2.5%) |
| Very low birth weight 1000-1499g | 9,136 (8.2%) | 16,436 (7.2%) | 1,643 (8.3%) | 6,352 (7.4%) | 33,567 (7.6%) |
| Low birth weight 1500-1999g | 15,202 (13.7%) | 29,327 (12.9%) | 2,063 (10.5%) | 11,911 (13.8%) | 58,503 (13.2%) |
| Low birth weight 2000-2499g | 15,322 (13.8%) | 40,418 (17.8%) | 2,091 (10.6%) | 13,936 (16.2%) | 71,767 (16.1%) |
| ≥2500g | 66,985 (60.2%) | 122,761 (54%) | 8,472 (43.0%) | 50,977 (59.2%) | 249,195 (56.1%) |
| Missing | 0 (0%) | 14,925 (6.6%) | 4,782 (24.2%) | 741 (0.9%) | 20,448 (4.6%) |
| Location of birth | | | | | |
| Born in the neonatal unit hospital (Inborn) | 86,855 (78.1%) | 173,494 (76.3%) | 9,910 (50.2%) | 56,323 (65.4%) | 326,582 (73.5%) |
| Born elsewhere (Outborn) | 16,951 (15.2%) | 48,559 (21.3%) | 9,797 (49.7%) | 29,423 (34.2%) | 104,730 (23.6%) |
| Missing | 7,418 (6.7%) | 5,466 (2.4%) | 16 (0.1%) | 368 (0.4%) | 13,268 (3%) |
| Discharge condition | | | | | |
| Alive | 95,598 (86%) | 194,657 (85.6%) | 16,384 (83.1%) | 74,566 (86.6%) | 381,205 (85.7%) |
| Dead | 15,529 (14%) | 30,528 (13.4%) | 3,323 (16.8%) | 11,058 (12.8%) | 60,438 (13.6%) |
| Missing | 97 (0.1%) | 2,334 (1.0%) | 16 (0.1%) | 490 (0.6%) | 2,937 (0.7%) |

**Appendix 1: Demographic characteristics of admitted neonates (January 2021–June 2025).**

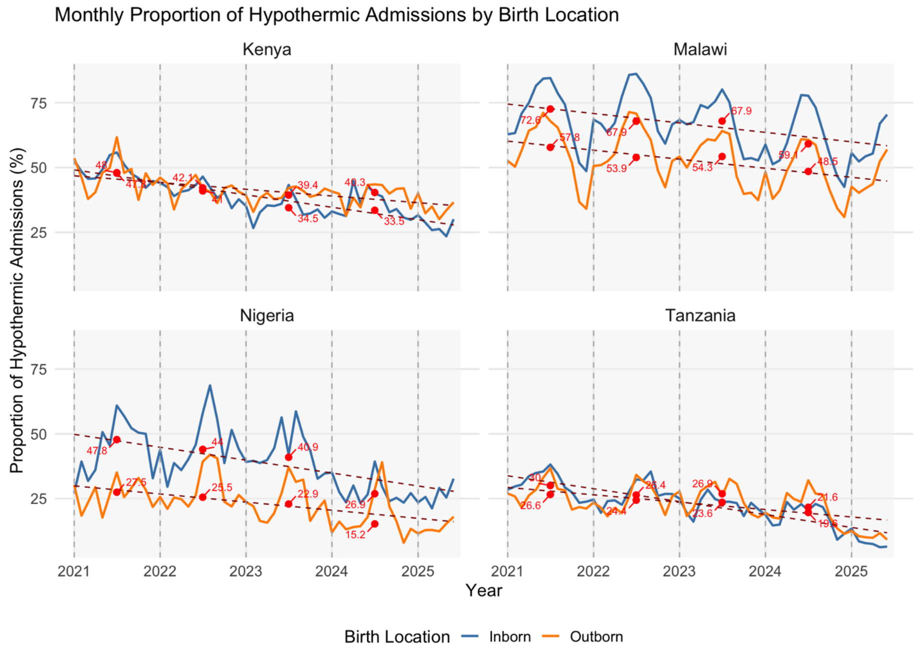


**Appendix 2: Monthly proportion of hypothermic admissions by birth location at the country level across years (January 2021–June 2025).** N=418,458.

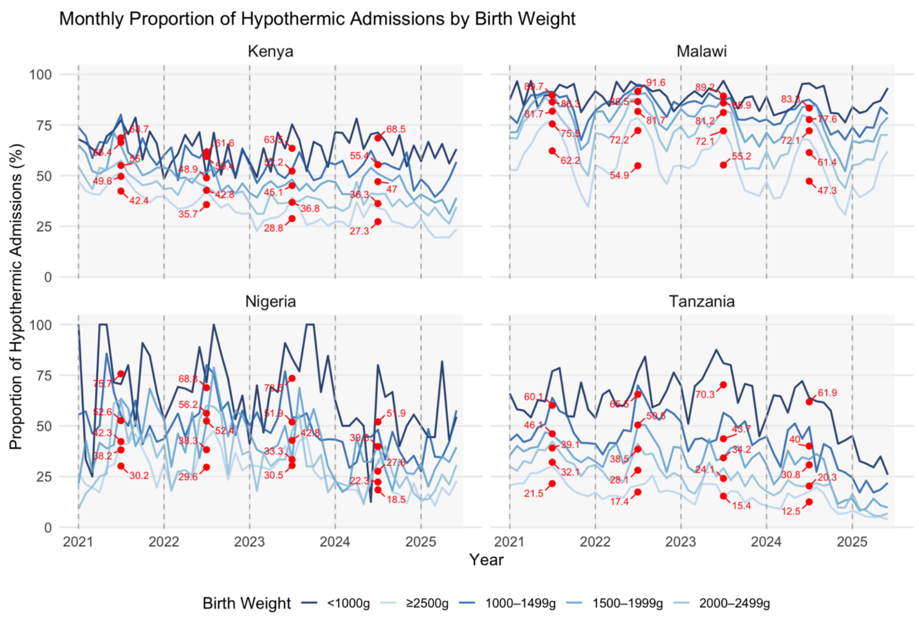


**Appendix 3: Monthly proportion of hypothermic admissions by birth weight at the country level across years (January 2021–June 2025).** N=418,458.

| | Kenya N (%) | Malawi N (%) | Nigeria N (%) | Tanzania N (%) |
|---|---|---|---|---|
| | N=9,539 | N=50,701 | N=3,811 | N=12,804 |
| Hypothermia at admission by birth location | | | | |
| Inborn | 2,118 (28.0) | 22,788 (59.0) | 554 (27.1) | 822 (9.3) |
| Outborn | 509 (33.7) | 5,011 (44.9) | 294 (16.7) | 431 (11.2) |
| Hypothermia after admission by birth location | | | | |
| Inborn | 4,998 (66.1) | 28,436 (73.7) | 1,126 (55.1) | 1,452 (16.4) |
| Outborn | 1,153 (76.4) | 8,576 (76.8) | 1,015 (57.7) | 910 (23.6) |
| Severity of hypothermia at admission | | | | |
| Mild hypothermia | 1,782 (18.7) | 8,953 (17.7) | 568 (14.9) | 839 (6.6) |
| Moderate hypothermia | 1,008 (10.6) | 19,063 (37.6) | 279 (7.3) | 408 (3.2) |
| Severe hypothermia | 7 (0.1) | 106 (0.2) | 1 (0.0) | 7 (0.1) |
| Severity of hypothermia after admission | | | | |
| Mild hypothermia | 3,913 (41.0) | 19,518 (38.5) | 1,501 (39.4) | 1,889 (14.8) |
| Moderate hypothermia | 2,567 (26.9) | 18,133 (35.8) | 643 (16.9) | 475 (3.7) |
| Severe hypothermia | 25 (0.3) | 73 (0.1) | 2 (0.1) | 7 (0.1) |

**Appendix 4: Thermal characteristics of admitted neonates, at and after admission (1 July 2024 to 30 June 2025) for all countries except Kenya (1 November 2024 to 30 June 2025).**

Monthly Neonatal Thermal Status Transitions in Malawi

50,701 complete records

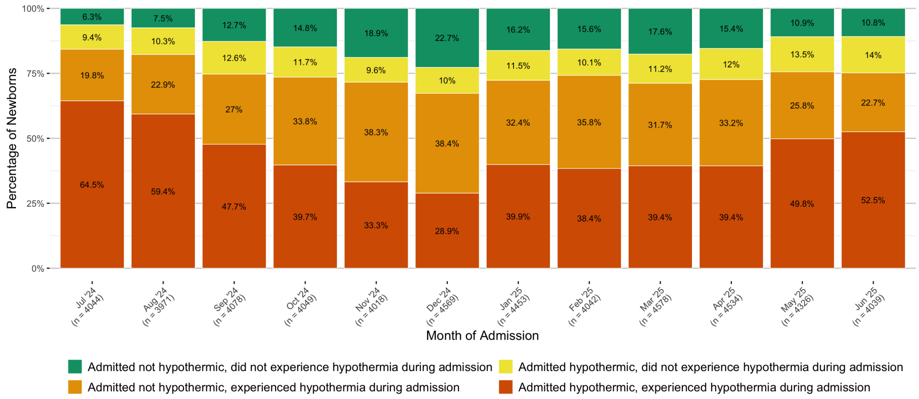


Monthly Neonatal Thermal Status Transitions in Nigeria

3,811 complete records

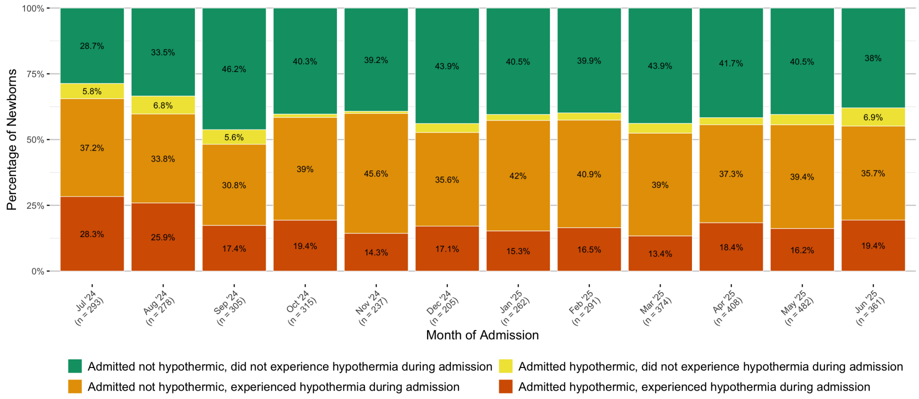

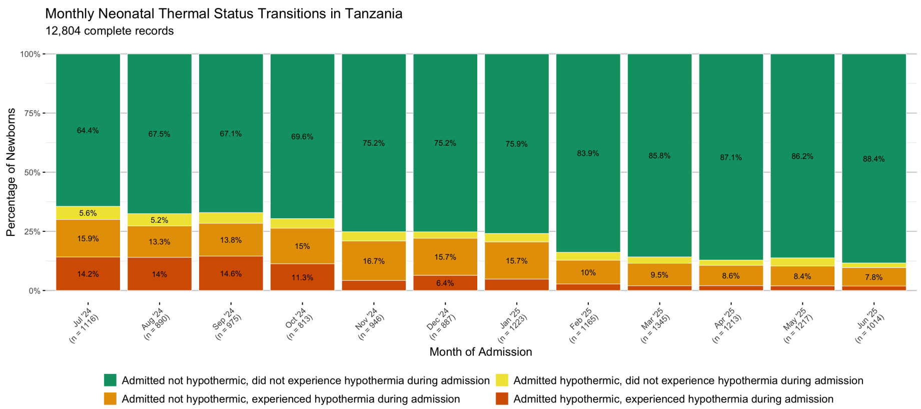


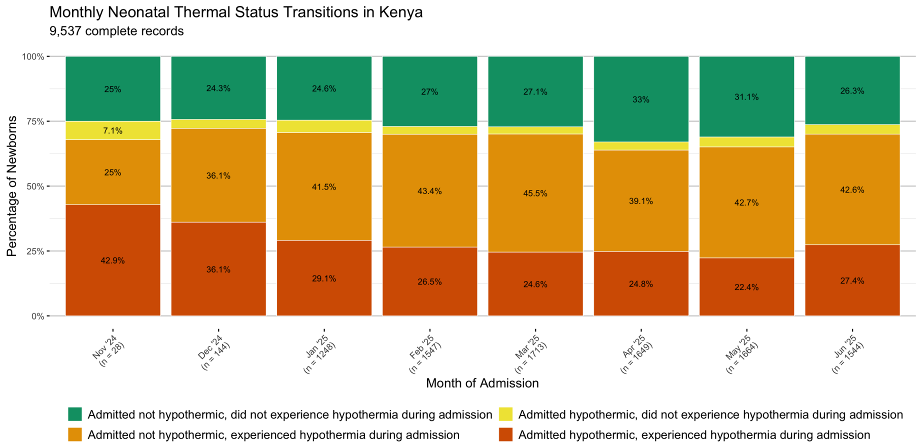


**Appendix 5: Country level neonatal thermal status transitions across 12 months of admission (1 July 2024 to 30 June 2025) except Kenya (1 November 2024 to 30 June 2025).**

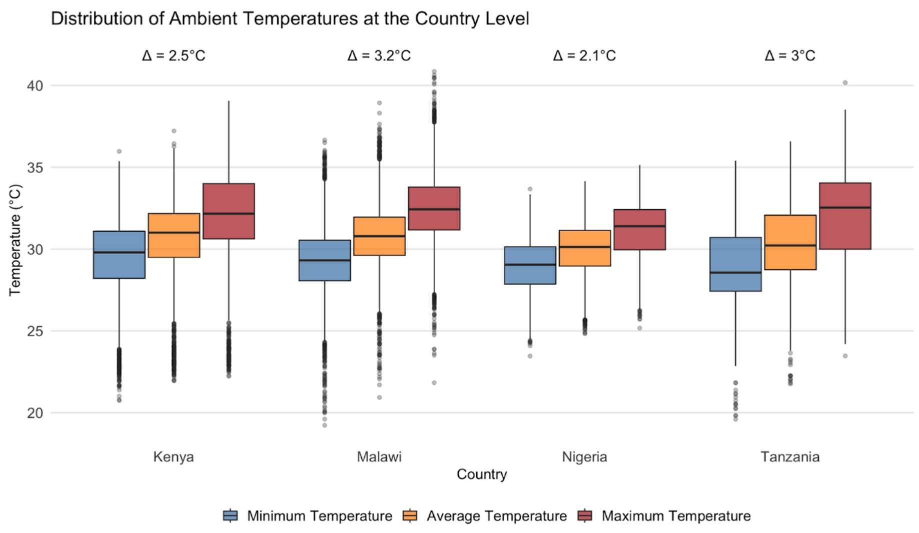


**Appendix 6: Distribution of minimum, average, and maximum ward temperatures at the country level from (1 July 2024 to 30 June 2025).** The average difference between maximum and minimum temperatures is annotated by country.

**Appendix 7: STROBE Checklist**

STROBE Statement—Checklist of items that should be included in reports of ***cross-sectional studies***

| | Item No | Recommendation | Page No, Line No |
|---|---|---|---|
| **Title and abstract** | 1 | (*a*) Indicate the study's design with a commonly used term in the title or the abstract | 2, 30 |
| | | (*b*) Provide in the abstract an informative and balanced summary of what was done and what was found | 2-3, 30-57 |
| **Introduction** | | | |
| Background/rationale | 2 | Explain the scientific background and rationale for the investigation being reported | 6-7, 61-99 |
| Objectives | 3 | State specific objectives, including any prespecified hypotheses | 9-10, 128-174 |
| **Methods** | | | |

| | | | |
|---|---|---|---|
| Study design | 4 | Present key elements of study design early in the paper | 8, 101-112 |
| Setting | 5 | Describe the setting, locations, and relevant dates, including periods of recruitment, exposure, follow-up, and data collection | 11, 115-118 and 175-184 |
| Participants | 6 | (*a*) Give the eligibility criteria, and the sources and methods of selection of participants | 11, 175-184 |
| Variables | 7 | Clearly define all outcomes, exposures, predictors, potential confounders, and effect modifiers. Give diagnostic criteria, if applicable | 9, 129-137<br>20, Table 3<br>21, Table 4 |
| Data sources/ measurement | 8* | For each variable of interest, give sources of data and details of methods of assessment (measurement). Describe comparability of assessment methods if there is more than one group | 8, 114-124 |
| Bias | 9 | Describe any efforts to address potential sources of bias | 10, 160-167<br>25, 447-454 |
| Study size | 10 | Explain how the study size was arrived at | 11, 175-184 |
| Quantitative variables | 11 | Explain how quantitative variables were handled in the analyses. If applicable, describe which groupings were chosen and why | 9-10, 128-174 |
| Statistical methods | 12 | (*a*) Describe all statistical methods, including those used to control for confounding | 9-10, 128-174 |
| | | (*b*) Describe any methods used to examine subgroups and interactions | N/A |
| | | (*c*) Explain how missing data were addressed | 9, 125-127 |
| | | (*d*) If applicable, describe analytical methods taking account of sampling strategy | N/A |
| | | (*e*) Describe any sensitivity analyses | 10: 160-167 |
| **Results** | | | |
| Participants | 13* | (a) Report numbers of individuals at each stage of study—eg numbers potentially eligible, examined for eligibility, confirmed eligible, included in the study, completing follow-up, and analysed | 11, 175-184 |
| | | (b) Give reasons for non-participation at each stage | 11, 175-184 |
| | | (c) Consider use of a flow diagram | N/A |

| | | | |
|---|---|---|---|
| Descriptive data | 14* | (a) Give characteristics of study participants (eg demographic, clinical, social) and information on exposures and potential confounders | 33, Appendix 1 |
| | | (b) Indicate number of participants with missing data for each variable of interest | 33, Appendix 1<br>9, 125-127 |
| Outcome data | 15* | Report numbers of outcome events or summary measures | 11, 186-189<br>12, 206-209 |
| Main results | 16 | (*a*) Give unadjusted estimates and, if applicable, confounder-adjusted estimates and their precision (eg, 95% confidence interval). Make clear which confounders were adjusted for and why they were included | 20, Table 3 |
| | | (*b*) Report category boundaries when continuous variables were categorized | 20, Table 3 |
| | | (*c*) If relevant, consider translating estimates of relative risk into absolute risk for a meaningful time period | N/A |
| Other analyses | 17 | Report other analyses done—eg analyses of subgroups and interactions, and sensitivity analyses | N/A |
| **Discussion** | | | |
| Key results | 18 | Summarise key results with reference to study objectives | 11-21, 175-341 |
| Limitations | 19 | Discuss limitations of the study, taking into account sources of potential bias or imprecision. Discuss both direction and magnitude of any potential bias | 25, 447-454 |
| Interpretation | 20 | Give a cautious overall interpretation of results considering objectives, limitations, multiplicity of analyses, results from similar studies, and other relevant evidence | 21-25, 342-435 |
| Generalisability | 21 | Discuss the generalisability (external validity) of the study results | 25, 441-443 |
| **Other information** | | | |
| Funding | 22 | Give the source of funding and the role of the funders for the present study and, if applicable, for the original study on which the present article is based | 27-28, 503-508 |

*Give information separately for exposed and unexposed groups.

**Commented [CB1]:** also reference same statement added above for 12 c

**Note:** An Explanation and Elaboration article discusses each checklist item and gives methodological background and published examples of transparent reporting. The STROBE checklist is best used in conjunction with this article (freely available on the Web sites of PLoS Medicine at http://www.plosmedicine.org/, Annals of Internal Medicine at http://www.annals.org/, and Epidemiology at http://www.epidem.com/). Information on the STROBE Initiative is available at www.strobe-statement.org.
